# SARS-CoV-2 seroprevalence and clinical features of COVID-19 in a German liver transplant recipient cohort: a prospective serosurvey study

**DOI:** 10.1101/2020.09.30.20204537

**Authors:** Conrad Rauber, Shilpa Tiwari-Heckler, Jan Pfeiffenberger, Arianeb Mehrabi, Frederike Lund, Philip Gath, Markus Mieth, Uta Merle, Christian Rupp

## Abstract

In liver transplant (LT) recipients with severe COVID-19 fatal outcome has been reported in a substantial subset of patients. Whether LT recipients are at increased risk for severe COVID-19 compared to the general population is controversial. Here we report the first results of a SARS-CoV-2 serosurvey in a large LT recipient cohort.

Taking into account known risk factors, LT recipients a priori represented a high-risk cohort for severe COVID-19 with 101/219 (46.1 %) presenting with more than 2 risk factors for severe COVID-19. Out of 219 LT recipients 8 (3.7%) were either tested positive for nasopharyngeal SARS-CoV-2 RNA or anti-SARS-CoV-2 serum IgG. 5/8 (62.5 %) did not show any clinical signs of infection, 3/8 (37.5%) had self-limited disease, none required hospitalization for COVID-19. 5/8 (67.5%) SARS-CoV-2 positive patients showed high utilization of the healthcare system. 2/8 (25 %) had known exposure to infected health care personal. A majority of 65.4 % often or always avoided outside family social contacts. Face masks were commonly worn by all patients.

In summary, LT recipients showed a SARS-CoV-2 seroconversion rate similar to the general population with a substantial percentage of unrecognized infections. The health care system can be the assumed source of infection in most of these cases.

## Introduction

Liver transplant (LT) recipients are a vulnerable patient group for severe infections due to immunosuppression. Two recently published case series in liver transplant and solid organ transplantation (SOT) recipients with COVID-19 reported a high fatality rate of up to 30% exceeding the rate of the general population.^1–4^ In contrast, a swiss case study found mortality rates comparable to comorbidity matched non-transplant recipients.^5^ Initial symptoms of COVID-19 patients are diverse.^6^ Patients who tested positive for SARS-CoV2 can be asymptomatic or can develop COVID-19 and in severe cases may deteriorate due to acute respiratory distress syndrome with subsequent respiratory failure and death.^7^ Both among the general population and SOT recipients the main risk factors for severe COVID-19 were older age and comorbidities like diabetes, obesity, renal and cardiopulmonary disease.^8–11^ Data on immunosuppression as a risk factor for COVID-19 is inconclusive, but minimization of immunosuppression (IS) has been widely recommended for LT patients with COVID‐19 based on experience with other respiratory pathogens.^12^ LT patients in most centers are therefore counselled to strictly follow hygiene and lifestyle restrictions to avoid SARS-CoV-2 infection.^13^ It is not known to which extend this adv is effective to reduce SARS-CoV-2 infection in the LT recipient population. This study aims to define incidence and prevalence of SARS-CoV-2 infection and to explore the (preventive) effect of personal behavioural adjustments of LT recipients.

## Methods

### Study design

This prospective study was conducted between March 5th, 2020 and August 6th, 2020 at the University Hospital Heidelberg located in the German Bundesland of Baden-Württemberg. All study participants had undergone liver transplantation in the past and were older than 18 years. All study participants provided written and informed consent before enrolment. The study was approved by the Ethics Committee of the Medical Faculty of the University of Heidelberg (approval number S-457/2020). The study was conducted in accordance with good clinical practice (GCP) and the Declaration of Helsinki.

### Sampling and procedures

Study enrollment was performed when patients presented for scheduled routine follow up after liver transplantation. Routine follow up on an outpatient basis is performed at least every six months at our center. After having provided written and informed consent, study participants completed a questionnaire querying information including demographics and behavior in the pandemic. Furthermore, study participants were asked to provide blood specimens and nasopharyngeal swabs. Blood was centrifuged and plasma was stored until analysis (−80°C). Analyses were performed in batches at the central laboratory of the University Hospital Heidelberg Infectious Disease department. Anti-SARS-CoV-2 IgG (EUROIMMUN) were determined with enzyme-linked immunosorbent assays. The data sheet (April 29, 2020) reports cross-reactivities with anti-SARS-CoV-1-IgG-antibodies, but not with MERS-CoV-, HCoV-229E-, HCoV-NL63-, HCoV-HKU1-or HCoV-OC43-IgG antibodies.

### Statistics

Statistical analysis was carried out by a statistician using Graph Pad (Version 4.4.1), SPSS (Version 22) as appropriate. P values are given as indicated using chi-square test or non-parametric t-test.

## Results

We conducted a prospective study of 219 liver transplant recipients at a liver transplant center in the south of Germany with sample collection between may 5^th^ and august 6^th^ 2020.

Liver transplant (LT) patients are a highly comorbid patient group. In our study only 55/219 (25.1 %) had no known risk factor for a severe course of COVID-19 (diabetes, hypertension, obesity, cardiac disease, leukopenia), while 63/219 (28.8 %) had 1 risk factor and 101/219 (46.1 %) had 2 or more risk factors. Particularly, arterial hypertension (108/219, 49.3 %) and type II diabetes mellitus (66/219, 30.1 %) were highly prevalent among our study participants (**table 1**). Routine pharyngeal and nasal SARS-CoV-2 swabbing was performed on 126 patients between may 5^th^ 2020 and July 5^th^ 2020. Thereafter routine SARS-CoV-2 screening was terminated because general population infection rates were very low. 2/136 (1.5 %) patients tested positive for nasopharyngeal SARS-CoV-2 RNA (**table 2**). Both SARS-CoV-2 RNA positive patients (patient 1 and 2, **table 3**) presented to planned routine LT follow up meetings but reported symptoms of uncomplicated upper respiratory tract infection.

**Table 1.**

|  | All patients<br>(n=219) | SARS-CoV-2<br>positive patients<br>(n=8) | p-value |
| --- | --- | --- | --- |
| **Sex, female (%)** | 89 (40.6%) |  |  |
| **Age, years [range]** | 56.9 (18.1-78.2) | 56.7 (25.5-69.5) |  |
| **>65 years** | 44 (20.8 %) | 2 (25.0%) |  |
| **Interval from liver,**<br>**years** | 6.4 (0.1-30.0) | 6.5 (0.2-15.0) |  |
| **Indication for LT** |  |  |  |
| **Alcohol** | 42 (19.2%) | 0 (0.0%) |  |
| **Hepatitis B** | 9 (4.1%) | 1 (12.5%) |  |
| **Hepatitis C** | 10 (4.6%) | 0 (0.0%) |  |
| **HCC** | 43 (19.6%) | 2 (25.0%) |  |
| **PSC** | 26 (11.9%) | 0 (0.0%) |  |
| **Others** | 79 (36%) | 5 (62.5%) |  |
| **Comorbidities** |  |  |  |
| **BMI, kg/m<sup>2</sup>** | 25.6 (15.8-46.0) | 22.9 (20.0-32.2) |  |
| **BMI<25** | 97 (44.3%) | 5 (62.5%) |  |
| **BMI 25-30** | 74 (33.8%) | 1 (12.5%) |  |
| **BMI >30** | 48 (21.9%) | 2 (25%) |  |
| **Cardiovascular**<br>**disease** | 31 (14.2%) |  |  |
| **Respiratory**<br>**disease** | 21 (9.6%) | 0 (0.0%) |  |
| **Hypertension** | 108 (49.3%) | 5 (62.5%) |  |
| **Diabetes** | 66 (30.1%) | 2 (25.0%) |  |
| **Active cancer** | 3 (1.4%) | 0 (0.0%) |  |
| **Active smoker** | 19 (8.7%) | 0 (0.0%) |  |
| **Leukopenia** | 50 (22.8%) | 2 (25%) |  |
| **RAAS inhibitor use** | 56 (25.6%) | 1 (12.5) |  |
| **Renal insufficiency** |  |  |  |
| **Dialysis** | 11 (5.0%) | 2 (25%) | 0.02 |
| **GFR<30ml/min** | 17 (7.7%) | 2 (25%) |  |
| **GFR 30-60ml/min** | 43 (19.6%) | 2 (25%) |  |
| **GFR<60ml/min** | 159 (72.6%) | 4 (50%) |  |
| **Immunosuppression** |  |  |  |
| **Tacrolimus** | 140 (63.9%) | 5 (62.5%) |  |
| **Ciclosporin** | 42 (19.2%) | 2 (25%) |  |
| **mTOR inhibitor** | 54 (24.7%) | 2 (25%) |  |
| **MMF/MPA** | 97 (44.3%) | 4 (50%) |  |

**Table 2.**

|  |  | Anti-SARS-CoV-2-IgG |  |  |
| --- | --- | --- | --- | --- |
|  |  | negative | positive |  |
| SARS-CoV-2<br>-RNA | Negative | 211 (96.3%) | 6 (2.7%) | 217 (99.0%) |
|  | positive | 1 (0.5%) | 1 (0.5%) | 2 (1.0%) |
|  |  | 212 (96.8%) | 7 (3.2%) |  |

**Table 3.**

|  | Sex | RNA | IgG | Age (years) | Time since LT (years) | Clinical symptoms | Dialysis | Hypertension | Diabetes | BMI | Immunosuppression |
| --- | --- | --- | --- | --- | --- | --- | --- | --- | --- | --- | --- |
| #1 | F | + | + | 20-30 | <1 | fever, dry cough, fatigue | No | No | No | 23 | Tac/MMF |
| #2 | F | + | - | 50-60 | 5-10 | dry cough | Yes | Yes | No | 30 | Cic |
| #3 | M | - | + | 60-70 | <1 | None | No | Yes | Yes | 30 | Tac/MMF |
| #4 | M | - | + | 60-70 | 1-5 | None | No | Yes | Yes | 32 | Tac/mTOR |
| #5 | F | - | + | 50-60 | 5-10 | None | No | Yes | No | 22 | Tac/MMF |
| #6 | M | - | + | 60-70 | 5-10 | None | No | No | No | 20 | Tac/MMF |
| #7 | M | - | + | 50-60 | 10-20 | None | No | No | No | 23 | Tac |
| #8 | F | - | + | 60-70 | 10-20 | None | Yes | Yes | No | 21 | mTOR |

We further collected 241 serum samples from 219 LT recipients presenting to routine follow up between may 5^th^ and august 6^th^ 2020 in southern Germany (**figure 1**). We performed a semi-quantitative ELISA on anti-SARS-CoV-2-IgG directed against its spike protein. Of these 241 samples 13 (5.4 %) tested positive for anti-SARS-CoV-2-IgG representing 7 individual patients. Some patients had repeat serum samples and all of them were concordantly positive or negative for anti-SARS-CoV-2-IgG. Overall seroprevalence among our study cohort for anti-SARS-CoV-2-IgG was 3.2 % (**table 2**). One of the patients that had tested positive for SARS-CoV-2-RNA tested negative for anti-SARS-CoV-2-IgG (**table 3**, patient 2). All patients who tested positive (IgG or RNA) were contacted on 10^th^ august and retrospectively surveyed for clinical symptoms of COVID-19 in the prior 6 months. Overall, only 3 patients (including the two patients we had identified by nasopharyngeal swabbing) recalled symptoms of flu like upper respiratory tract infection possibly related to COVID-19, while 5 did not (3/8, 37.5 %). For two patients we had temporally distinct serum samples available. Both showed a steep decline in antibody titers over time (**figure 2**). One patient that tested strongly positive for anti-SARS-CoV-2-IgG upon diagnosis was formally seronegative by the end of the study (**figure 2 & table 3**, patient 1). All three clinically apparent COVID-19 cases were mild and self-limited. Patient 1 however showed severe blood test abnormalities with severe thrombocytosis, coagulopathy and elevated heart muscle enzymes.

**Figure 1.**
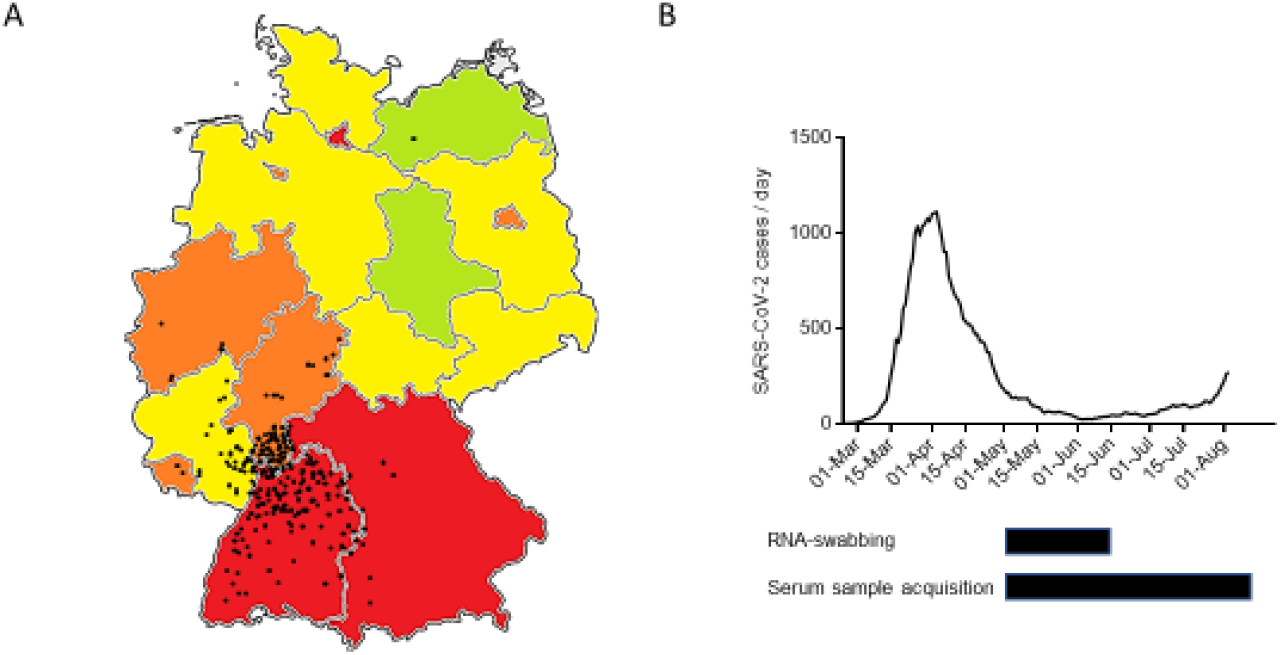
The course of the SARS-CoV-2 pandemic in Germany **A** Map of the federal republic of Germany indicating RNA proven SARS-CoV-2 cases per capita in different federal states. Red > 300 cases /100,000, Orange 200-300 cases/100,00, yellow 100-200cases/100,000, green <100 cases/100,000 as of August 15th. Black dots indicate residence of enrolled LT recipients. B Number (7-days gliding average) of RNA proven SARS-CoV-2 cases per day in the federal state of Baden-Württemberg from february 25th until august 8th. Below sample accrual time frame for nasopharyngeal RNA swab (upper black bar) and blood serum sample (lower black bar).

**Figure 2.**
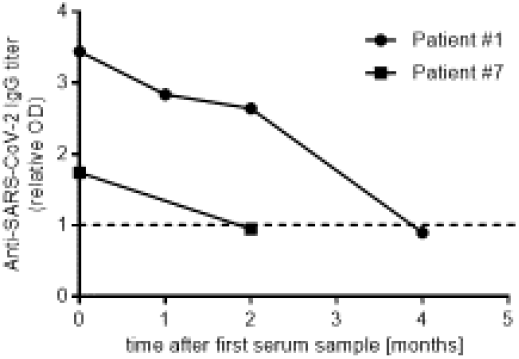
Anti-SARS-CoV-2 serum titers in patients #1 and #7 over time.

Of the seroconverted patients 2/8 had potential contact with infected health care personal during a hospital stay. For the others the source of infection was unknown. Interestingly, 2/8 were recently transplanted, 2/8 were on dialysis for chronic renal failure and 1/8 lived in an assisted living community.

104 patients were surveyed at the time of sample collection for the use of protective equipment and personal behavior adjustments in the COVID-19 pandemic. 37/104 (35.6 %) were highly or very highly concerned about contracting SARS-CoV-2, while 31/104 (29.8 %) felt the pandemic highly or very highly interfered with their daily lives. 68/104 (65.4 %) often or always avoided outside family social contacts and 66/104 (63.5 %) often or always avoided going to supermarkets or shops, but only 43/104 (41.3 %) often or always avoided medical visits (**figure 3, A-F**). Most commonly worn face masks were surgical masks (47.5 %), cloth masks (31.7 %) and FFP-2 masks (20.8 %) (**figure 3, G**). 100% always wore masks in supermarkets and in hospitals and 16.3 % always wore masks outside.

**Figure 3.**
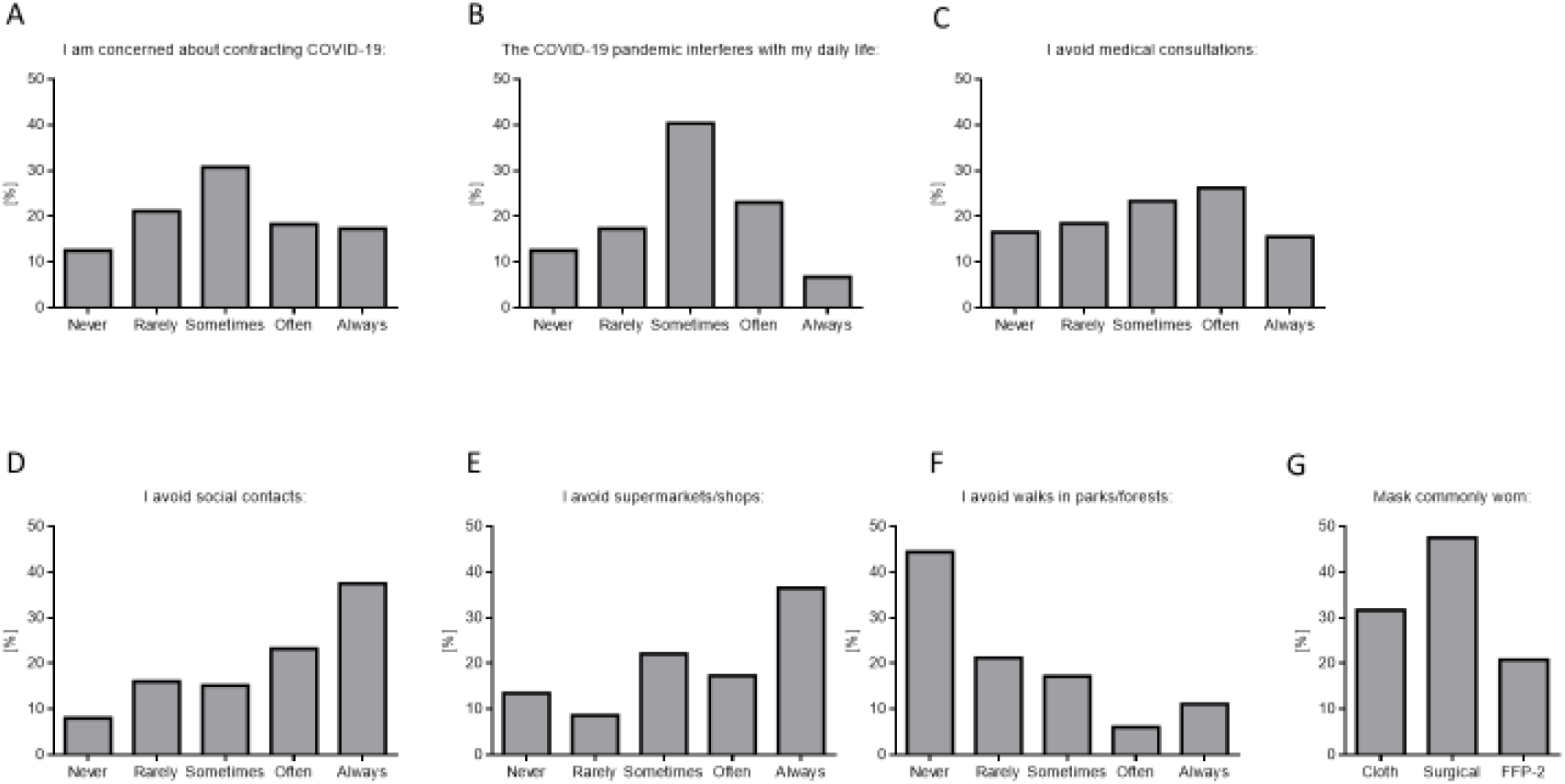
Survey of self-protection measurements and behavioural adjustments among LT recipients during the SARS-CoV-2 pandemic.

## Discussion

We performed a prospective screening trial for SARS-CoV-2-RNA and anti-SARS-CoV-2-IgG infection in LT recipients during the COVID-19 pandemic in southern Germany. We documented acute or past SARS-CoV-2 infection in 3.7 % of our LT recipients during the study. While direct serosurveys in the general population are lacking for our study region, the SARS-CoV-2 infected percentage in the population can be roughly estimated from serosurvey studies in other regions with similar health care setting. Until study termination about 37.000 PCR proven SARS-CoV-2 infections had been documented in the study area (Baden-Württemberg). Recent German serosurveys provide a rough estimate of a factor 4 – 10 of PCR documented SARS-CoV-2 infections to overall serological proven infections proposing a seroprevalence of 1.3-3.2 % in south-western Germany.^14,15^ Taking into account this estimate, the prevalence of active or past SARS-CoV-2 infection in LT recipients is comparable to the general population. Our direct study was limited to 219 patients that presented for routine follow up appointments after liver transplantation at our institution. However, the overall cohort comprises 1200 patients in the same region. These patients were encouraged to contact the transplant center in case of infection or hospital stay. Over the study period we did not record a single case of hospitalization or death due to COVID-19 in this cohort. The presented data shows an overall low but significant percentage of LT patients that had been infected with SARS-CoV-2. 5/8 patients serovconverted without experiencing any clinical overt symptom, in the other 3/8 patients clinical symptoms were mild and self-limited Interestingly, patient 2 (table 3) screened positive for SARS-CoV-2-RNA, but was negative for anti-SARS-CoV-2-IgG on multiple occasions afterwards. It cannot be ruled out that we missed patients in our cohort, because they simply did not develop anti-SARS-CoV-2 antibodies. The detailed cases of patient 1 and 3 (**table 3**) with repetitive serum samples highlight the potential rapid loss of anti-SARS-CoV-2-IgG within few weeks. To what extend immunosuppression adds to the phenomenon could not be assessed due to the overall low case number. Most LT recipients were aware of the risk of SARS-CoV-2 infection and protected themselves with face masks and avoided public places such as supermarkets or social gatherings. While a control group is certainly missing, it is surprising that despite the self-containment efforts in this patient cohort the SARS-CoV-2-IgG seroprevalence was comparable to the general population. Examining detailed patient information highlights the role health care acquired infection might play. 5/8 patients were hospitalized during the pandemic or underwent hemodialysis for chronic renal failure. We cannot rule out that these patients contracted SARS-CoV-2 infection in the healthcare setting and that the healthcare system itself poses an important risk for these comorbid patient group. Concordantly, health care visits were less strictly avoided than supermarkets or social contact (**figure 3**). The study poses several important limitations. Firstly, overall recorded numbers of seroconversion in the LT recipient cohort remained low and while the specificity of the antibody test is given at 99,6% by the manufacturer, we cannot rule out false positives. However, all positive tests were in the high titer range. Secondly, the study was insufficiently sized to assess individual risk factors such as immunosuppression, and it remains an important research question whether immunosuppression exerts protective effects against severe COVID-19 in LT patients. Compared to other studies that have highlighted an increased morbidity and mortality in LT recipients with COVID-19, our study was conducted prospectively in a healthcare setting that was at no point overwhelmed during the first wave of the pandemic. Risk for severe COVID-19 in LT recipients seems comparable to the general population and COVID-19 morbidity was low in a large LT recipient cohort. However, only 3.7 % had antibodies against SARS-CoV-2 leaving potentially 96.3 % of the cohort susceptible to SARS-CoV-2 infection. We documented a fast decline of anti-SARS-CoV-2-antibodies in LT recipients, questioning the durability of anti-SARS-CoV-2 immunity and emphasizing the importance of continued containment efforts to protect patient groups at risk.

### Disclosure

The authors of this manuscript have nothing to disclose as described by the American Journal of Transplantation.

## Data Availability

Data is available upon request. contact

## Abbreviations

COVID-19: coronavirus disease 2019
CRP: C-reactive protein
LT: liver transplantation
PCR: polymerase chain reaction
SOT: solid organ transplantation
SARS-CoV-2: Severe acute respiratory syndrome coronavirus 2

